# Enhancing Kidney Failure Analysis: Web Application Development for Longitudinal Trajectory Clustering

**DOI:** 10.1101/2023.05.31.23290804

**Authors:** Yuan Gu, Yishu Gong, Mingyue Wang, Song Jiang, Chen Li, Zheng Yuan

## Abstract

Kidney failure is a critical health condition with significant impact on patient well-being and healthcare systems worldwide. Analyzing the longitudinal trajectory of kidney function is crucial for understanding disease progression, predicting outcomes, and personalizing treatment strategies. This paper proposes a novel approach utilizing latent longitudinal trajectory clustering techniques by incorporating survival information to analyze kidney failure and explore patterns within patient populations. Besides, we also developed a web application to provide visualize and intuitive way to explore the relationship between estimated glomerular filtration rate (EGFR) progression and survival outcomes, helping researchers and clinicians gain valuable insights. By identifying distinct subgroups, this analysis can aid in early detection, risk stratification, and treatment optimization. The proposed methodology holds promise for improving patient care and outcomes in the field of nephrology.

## 1. Introduction

Kidney failure, also known as end-stage renal disease (ESRD), represents a major health challenge globally. There are more than 1500 ESRD people per million population in countries with a high prevalence, such as the US [1]. Chronic kidney disease (CKD) is marked by slow and progressive decline in kidney function, leading to ESRD. The Global Burden of Disease (GBD) studies have revealed that chronic kidney disease (CKD) has emerged as a prominent contributor to global mortality [2]. CKD poses a significant clinical threat for two primary reasons. Firstly, renal impairment can serve as a precursor to the development of ESRD, which necessitates interventions such as dialysis and transplantation. Secondly, CKD amplifies the risk of cardiovascular complications, further exacerbating the potential health consequences associated with the condition [3]. Hence, gaining a comprehensive understanding of the progression of CKD is imperative and calls for urgent attention.

The diagnosis of CKD is typically established through laboratory testing, with a primary focus on estimating the glomerular filtration rate (EGFR) using filtration markers such as serum creatinine or cystatin C. These markers play a vital role in assessing kidney function and aiding in the diagnosis of CKD [4]. However, the determination of the duration for assessing CKD in accordance with the chronicity criterion lacks consensus among researchers and clinicians. Epidemiological studies employ diverse algorithms, ranging from single measurements to any repeated measurements beyond 90 days. Some studies limit the evaluation to measurements taken within a 90 to 365-day timeframe. Additionally, there is variation in the criteria used, with some studies requiring consecutive repeated markers of CKD, while others accept CKD markers interspersed with markers not conforming to CKD criteria. The absence of a standardized approach highlights the need for further research and consensus to establish a unified framework for assessing CKD duration [4, 5].

The majority of previous studies on CKD have predominantly been retrospective or cross-sectional in nature. Unfortunately, there is a scarcity of studies that have focused on investigating the long-term, time-varying disease progression and clinical risk factors associated with CKD. This knowledge gap underscores the critical need to develop robust longitudinal approaches for the diagnosis and prognosis of CKD patients [6-9]. By adopting a longitudinal perspective, researchers and clinicians can gain valuable insights into the dynamic nature of CKD, capturing the changes and patterns that occur over time. Such studies can help elucidate the trajectory of the disease, identify key factors that contribute to its progression, and uncover novel risk markers that may enhance prediction and prognosis. A longitudinal diagnosis and prognosis for CKD patients would significantly enhance clinical decision-making and patient management. It would enable healthcare professionals to assess the evolution of the disease, tailor treatment plans accordingly, and intervene proactively to mitigate adverse outcomes. Moreover, a deeper understanding of long-term disease progression and associated risk factors can pave the way for personalized medicine approaches, allowing for more precise and targeted interventions. Therefore, there is a pressing need to prioritize the development and implementation of longitudinal studies in CKD research.

On the other hand, there has been limited progress in the development of web applications that facilitate the presentation of how risk factors, such as EGFR, progress over time. The availability of a user-friendly online tool for doctors and clinical professionals would greatly enhance their ability to track and monitor the progression of risk factors associated with CKD. Such a tool could provide convenient access to longitudinal data, enabling real-time analysis and visualization of EGFR trends over time. The development of such an application holds great potential in improving clinical decision-making and facilitating more efficient patient management in the field of nephrology. Limited sources exist in the literature that address the development of web applications specifically related to CKD. For instance, a notable example is the work by Mariella Gregorich et al. [10], who developed a web application utilizing a prediction model for estimating future glomerular filtration rate in individuals with Type 2 diabetes and CKD. However, their model is based on a linear mixed-effects approach and does not incorporate survival information. In our previous study [11], we also contributed to the field by developing a web application that focuses on disclosing the risk profile for breast cancer. However, it is important to note that our application solely relies on time-to-event data and lacks a comprehensive longitudinal analysis component. The afore mentioned examples shed light on the scarcity of web applications that effectively integrate longitudinal analysis and survival information in the context of CKD. However, recognizing the dynamic nature of CKD progression and the impact of survival outcomes, it becomes imperative to develop more comprehensive web applications that encompass both longitudinal analysis techniques and survival information.

In our research, we addressed this gap by utilizing a latent mixed class model EGFR while incorporating survival Cox regression and considering other important covariates such as age, treatment group, and gender. This approach allows for a more comprehensive and accurate prediction of disease progression and outcomes in CKD patients. To facilitate the practical implementation of our research findings, we developed an R Shiny web application. This application provides an interactive platform that presents visualizations of the latent class trajectories, enabling healthcare professionals to better understand and interpret the complex patterns of disease progression. By integrating longitudinal analysis, survival information, and covariate effects, the web application equips healthcare professionals with valuable tools for making informed decisions, personalizing treatment plans, and ultimately enhancing patient care and management.

## 2. Data and Methods

The analysis conducted in our study utilized the MASTERPLAN dataset, which consisted of 505 patients with a baseline estimated glomerular filtration rate (denoted as EGFR in the following text) greater than 30 ml/min per 1.73m^2^. After excluding individuals under the age of 35, we had a final sample of 443 unique subjects for our analysis. The age range of the subjects was from 35.04 to 87.47 years old, and the EGFR range is from 15 to 102. Among the participants, there were a total of 305 males. The subjects were divided into two groups for comparison: the control group (n=222) and the Nurse Practitioner intervention group (n=221). The mean value of EGFR, calculated using the CKD-EPI Creatinine Equation, was found to be 48.405 ml/min per 1.73m^2^. Within the cohort, 47 subjects had reached CKD stage 5, indicating advanced kidney disease. These details provide essential information about the sample size, demographic characteristics, group distribution, and clinical parameters of the patients included in the analysis. The original dataset is available online to be downloaded in the following link: https://easy.dans.knaw.nl/ui/datasets/id/easy-dataset:124398/tab/2. Table 1 displays the characteristics of key variables by event group.

**Table 1.**
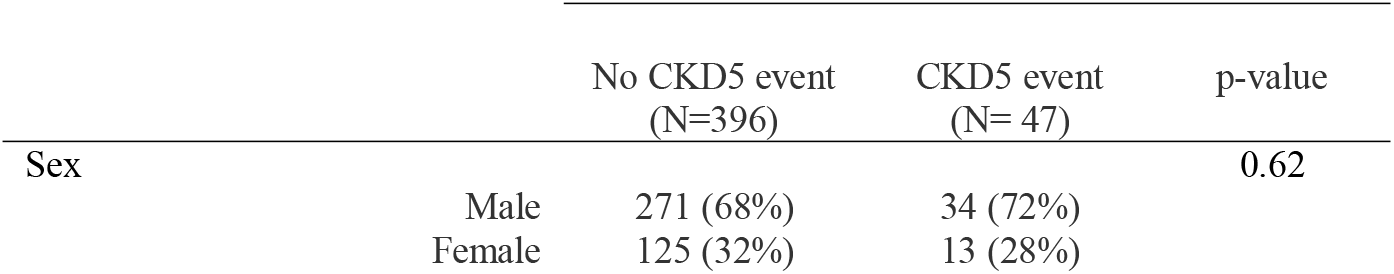
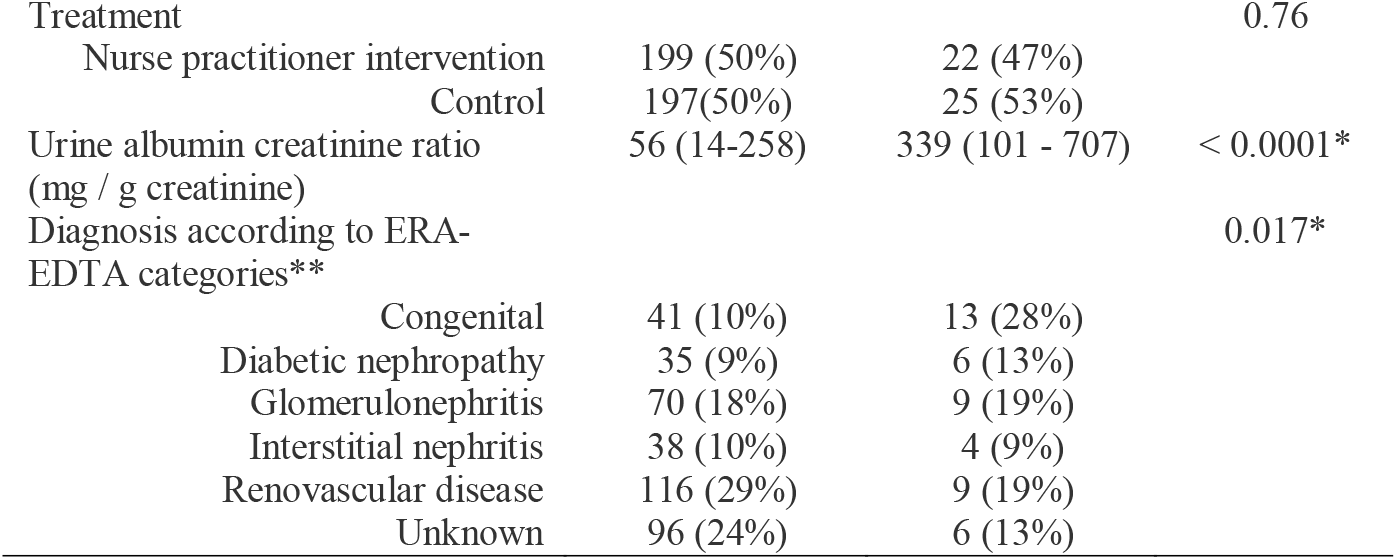
Descriptive statistics. * p value denotes significant level when p<0.05 ** continuous variables are summarized by mean (1st – 3rd Quantile)

|  |  | No CKD5 event<br>(N=396) | CKD5 event<br>(N= 47) | p-value |
| --- | --- | --- | --- | --- |
| Sex | Male | 271 (68%) | 34 (72%) | 0.62 |
|  | Female | 125 (32%) | 13 (28%) |  |

Table 1 Descriptive statistics. \* p value denotes significant level when $p < 0.05$
|  |  |  |  |
| --- | --- | --- | --- |
| Treatment |  |  | 0.76 |
| Nurse practitioner intervention | 199 (50%) | 22 (47%) |  |
| Control | 197(50%) | 25 (53%) |  |
| Urine albumin creatinine ratio<br>(mg / g creatinine) | 56 (14-258) | 339 (101 - 707) | < 0.0001* |
| Diagnosis according to ERA-EDTA categories** |  |  | 0.017* |
| Congenital | 41 (10%) | 13 (28%) |  |
| Diabetic nephropathy | 35 (9%) | 6 (13%) |  |
| Glomerulonephritis | 70 (18%) | 9 (19%) |  |
| Interstitial nephritis | 38 (10%) | 4 (9%) |  |
| Renovascular disease | 116 (29%) | 9 (19%) |  |
| Unknown | 96 (24%) | 6 (13%) |  |

### Longitudinal Trajectory Clustering

The Linear Mixed Model (LMM) [12] has emerged as a standard extension of classic statistical procedures, offering enhanced flexibility in the analysis of correlated longitudinal data. By allowing for the modeling of covariance structures that capture random effects, LMM enables the examination of changes over time in a more comprehensive manner. However, the analysis of longitudinal data can often be intricate and challenging, with the simple linear mixed model proving inadequate to capture the complex data structure and patterns. There are instances where the assumptions of the linear mixed model do not hold, such as when there is homogeneity in the populations being studied or when the longitudinal outcomes exhibit non-Gaussian distributions, including binary or ordinal responses. Hence, in our study the Latent Class Mixed Models (LCMM) is used.

The LCMM [13] combines the advantages of latent class analysis (LCA) and mixed effects models, allowing for the identification of unobserved subgroups or latent classes within the population while accounting for the correlation structure of repeated measurements over time. In LCMM, the data are assumed to arise from a mixture of latent classes, each representing a distinct subgroup with its own unique trajectory or pattern of change over time. These latent classes are unobserved or hidden variables that explain the underlying heterogeneity in the population. The following is the formular of the model:

We assume each one subject in the *N* subjects belongs to one unique class *g* of the *G* number of latent classes (*g =* 1*…G*). The latent class for subject *i* is noted as *c*_*i*_ =*g* when subject *i* belongs to class *g*:

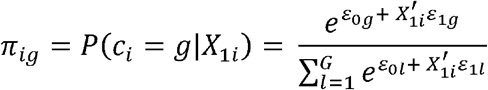

Repeated measures of the longitudinal marker *Y*_*ij*_ (*j* = 1, …, *n*_*i*_):

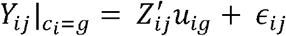

Where 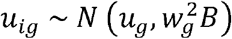 *and* ϵ_*ij*_ ∼ *N* (0, *σ*^2^)

However, there is no one criteria for optimizing the number of classes *G* due to different goals. To determine the number of latent classes, it often involves a balance between model fit, interpretability, and the specific research objectives. Exploratory analyses, sensitivity analyses, and expert input can aid in the decision-making process. For example, statistical selection criteria like Bayesian Information Criterion (BIC) or likelihood ratio tests can provide guidance. Subgroup sizes should also be considered, aiming for sufficient individuals in each class for meaningful analysis and interpretation, while avoiding excessive small sample sizes that lead to unstable estimates. Additionally, interpretability is also essential to assess if the identified latent classes make sense conceptually and align with prior knowledge or theory and represent distinct and interpretable subgroups.

### Log-rank test for latent classes

In our analysis, we integrate the visualization of survival differences obtained from log-rank tests as a criterion for selecting the number of latent classes. The log-rank test is commonly used in survival analysis to compare the survival distributions between different groups or classes. By incorporating the log-rank test results into the selection process, such as through Kaplan-Meier curves, we can examine the survival differences among the potential latent classes. This allows us to evaluate whether the identified latent classes exhibit distinct survival patterns or have statistically significant differences in survival outcomes.

In our analysis, the log-rank test does not directly involve the input of latent classes. Instead, it compares the observed number of events (such as deaths or failures) in each group to the expected number of events under the null hypothesis of no difference between the groups. The latent classes are determined separately through latent class modeling, and the log-rank test is used subsequently to assess the differences in survival patterns between the identified groups. The null hypothesis assumes that the survival functions of the latent groups being compared are identical. Suppose we have total *G* number of latent classes for longitudinal trajectories of the *N* subjects, he formula for the log-rank test statistic is as follows:

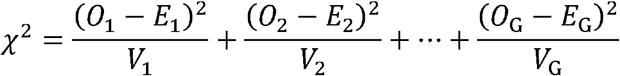

where:

- *O*_*i*_ represents the observed number of events in group.
- *E*_*i*_ represents the expected number of events in group *i* under the null hypothesis.
- *V*_*i*_ represents the variance of the observed number of events in group *i*.

The log-rank test statistic follows a chi-square distribution with degrees of freedom equal to *G-1*. By comparing the calculated test statistic to the critical value from the chi-square distribution, we can determine whether there are significant differences in survival outcomes among the groups.

## 3. Results and Findings

To facilitate the visualization of the analysis results and enhance understanding of the EGFR progression over time and its impact on survival rates, we have developed an interactive R Shiny web application. This web application provides a user-friendly interface and can be accessed via the following link: https://baran-shad.shinyapps.io/Kidney_failure/.

Pictures 1 and 2 illustrate the trajectory trends and corresponding survival differences when the number of latent classes is set to 3. In Picture 1, we observe distinct patterns among the latent classes. Class 1 (red curve, 96 subjects) demonstrates consistently high EGFR values around 75, with a slight decreasing trend as age increases. Class 2 (green curve, 333 subjects) exhibits relatively low EGFR values around 35, remaining relatively stable across different age groups. Class 3 (blue curve, 14 subjects) displays an inverted U-shaped curve, indicating an initial increase in EGFR values from 50 to 100 before age 60, followed by a declining trend decreasing to below 25 after age 60. Picture 2 shows that Class 1 has the highest survival rate, indicating a more favorable prognosis for individuals in this group. Conversely, Class 3 exhibits the lowest survival rate, suggesting a higher risk or poorer prognosis for individuals belonging to this class. In Picture 3, we observe that Class 1 and Class 4 (red and purple curves, 69 and 25 subjects respectively) demonstrates consistently high EGFR values around 75, although both have a slight decreasing trend as age increases after age 70, the two groups have highest survival rate in Picture 4. While Class 3 (blue curve, 131 subjects) exhibits relatively constant EGFR values above 50, remaining relatively stable across different age groups, it also has a high survival rate in Picture 4. In contrast, Class 2 (yellow curve, 218 subjects) displays constantly lower value of EGFR below 30 over all age groups and has the lowest survival rate.

**Picture 1:**
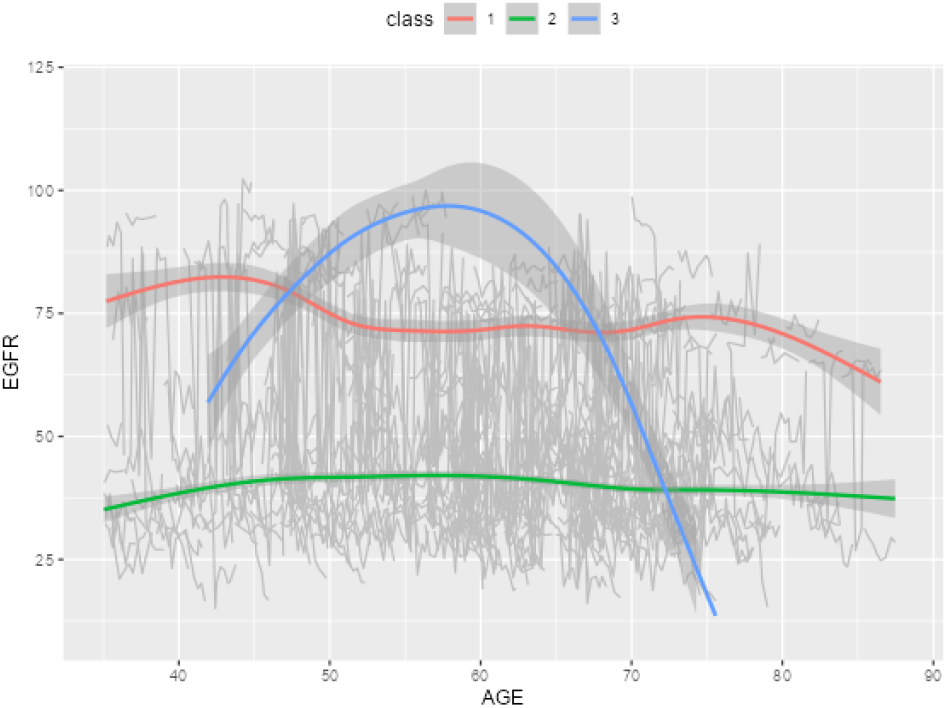
EGFR trajectory trends of the 3 latent classes over age.

**Picture 2.**
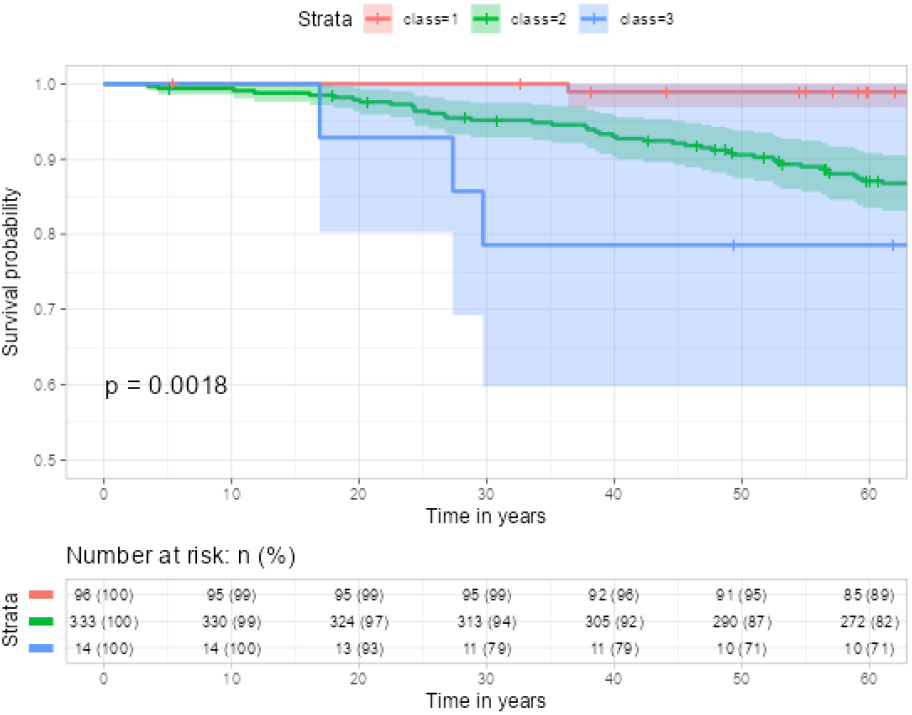
Survival difference of the 3 latent classes.

**Picture 3:**
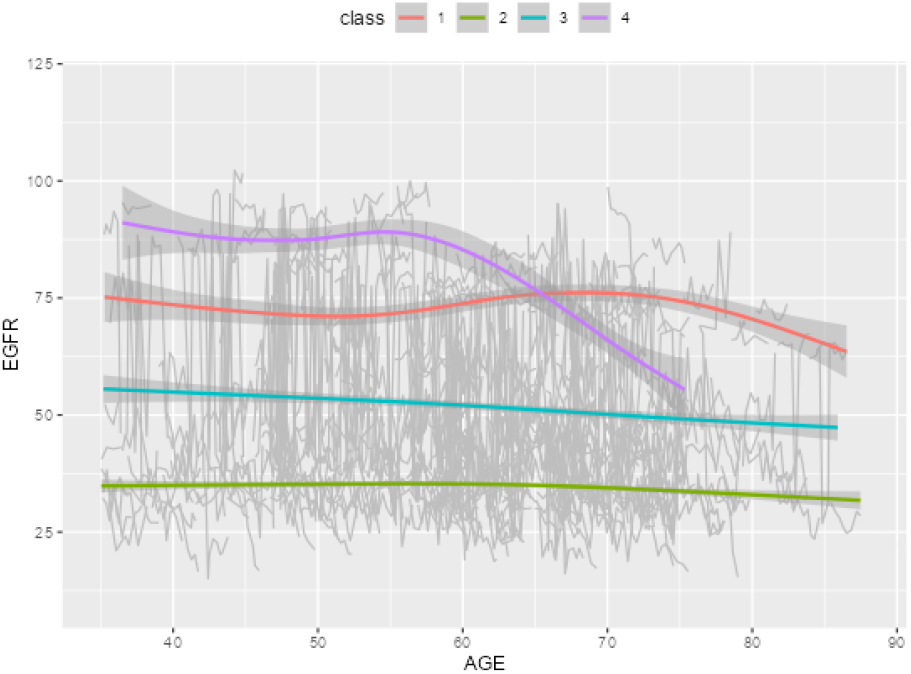
EGFR trajectory trends of the 4 latent classes over age.

**Picture 4:**
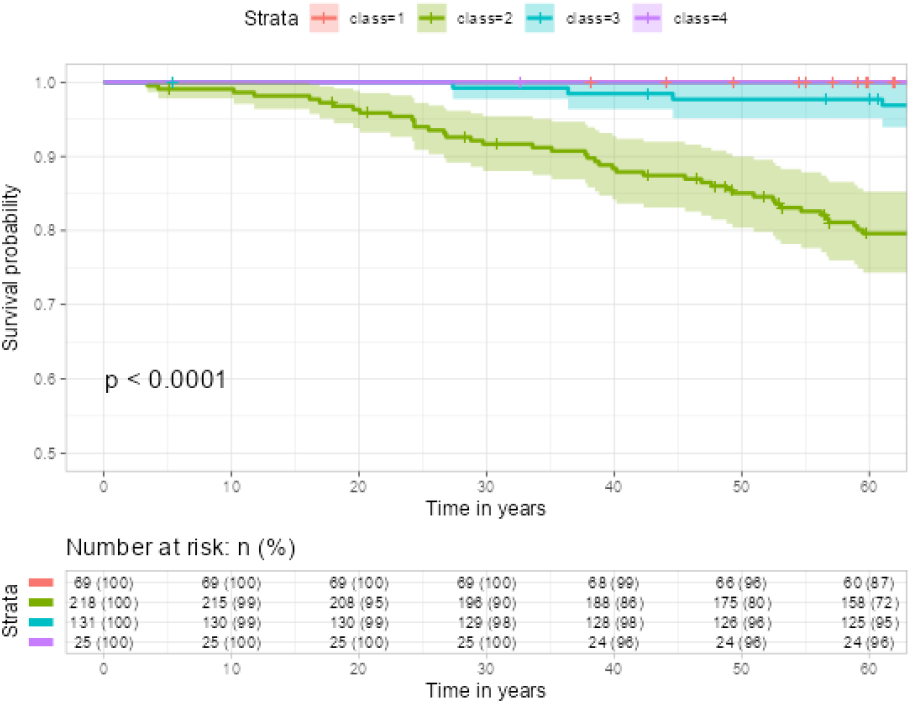
Survival difference of the 4 latent classes.

In Picture 5 and 6, we observe the trajectory patterns of different classes. Class 1 (red curve, 160 subjects) demonstrates consistently low EGFR values before age 60, followed by a rapid increase after age 60. This class has the lowest survival rate. Class 4 (blue curve, 42 subjects) exhibits a decreasing trend in EGFR values from 90 to below 25 throughout all ages, resulting in the second lowest survival rate. Class 3 (green curve, 138 subjects) also shows a decreasing EGFR trend, but less steeply compared to Class 4. The EGFR values range from 60 to 25, and this class has the third lowest survival rate. In contrast, Class 2, and Class 5 both display high EGFR values across all ages and have high survival rates. All p-values of the log rank tests are < 0.05. When incorporating latent class as an input covariate along with other covariates such as Diagnosis (DX) according to ERA-EDTA categories and Urine albumin creatinine ratio (UACR), the survival models achieved concordance values of 0.787, 0.728, and 0.718 for cluster numbers 3, 4, and 5, respectively.

**Picture 5:**
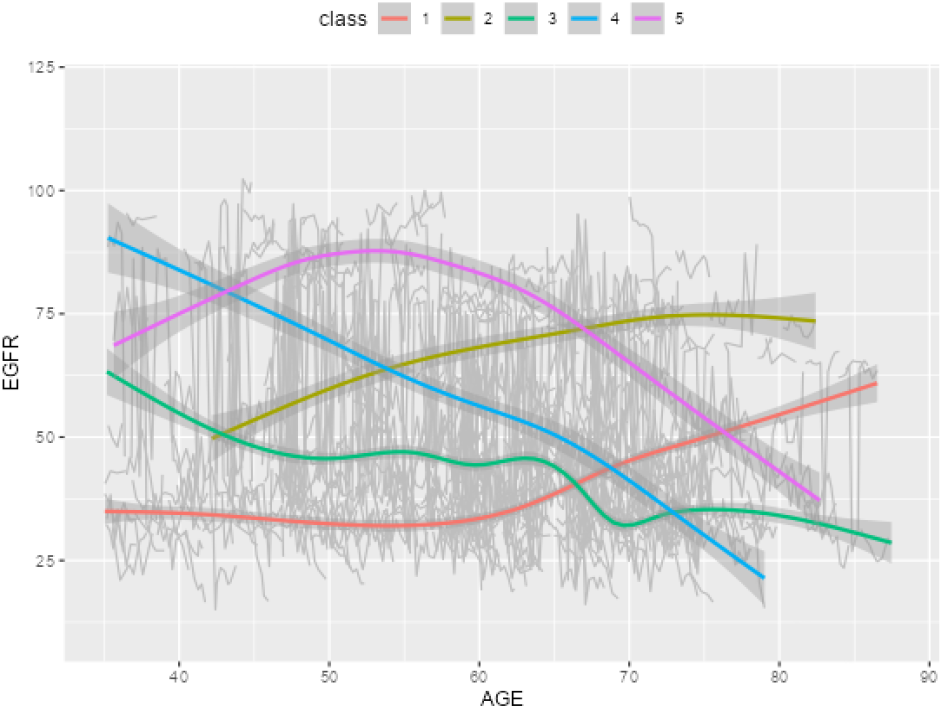
EGFR trajectory trends of the 5 latent classes over age.

**Picture 6:**
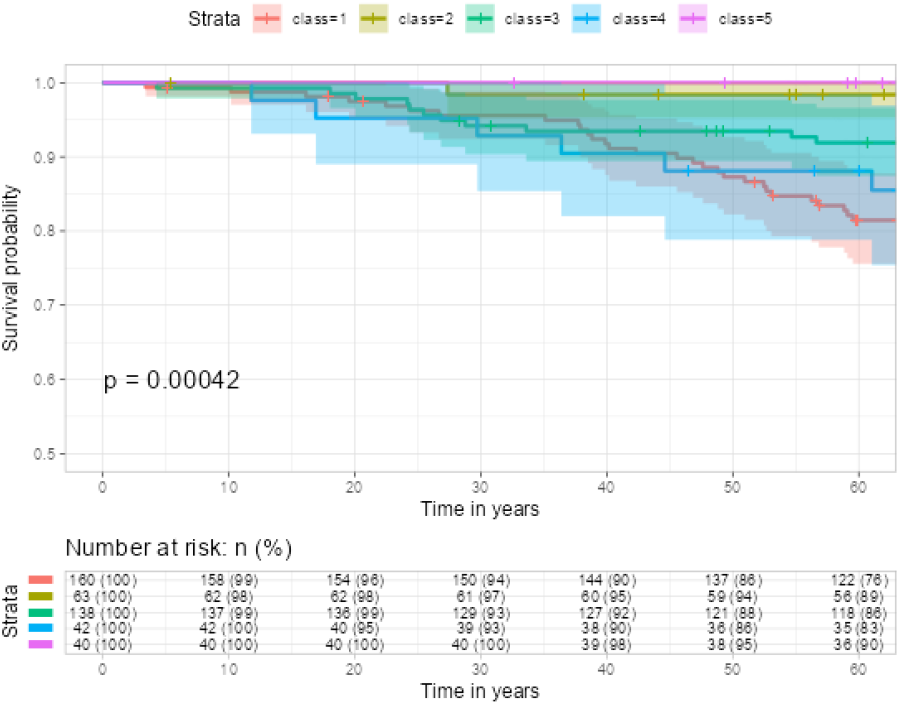
Survival difference of the 5 latent classes.

## 4. Conclusion

Our integrated approach combining latent longitudinal trajectory classes and survival analysis, presented through an online web application, has revealed insightful findings. Individuals with consistently low or rapidly declining EGFR values below 25 exhibit a higher risk for CKD5 events and lower survival rates compared to other groups. These results provide valuable knowledge for risk stratification and personalized care in kidney disease.

The findings include the identification of distinct subgroups within the patient population based on their kidney function trajectories. The paper discusses the characteristics and clinical relevance of these subgroups, highlighting any associations with disease progression, treatment response, or patient outcomes. This interactive platform provides an intuitive way to explore the relationship between EGFR progression and survival outcomes, helping researchers and clinicians gain valuable insights.

However, it is important to note that the current analysis is limited by the availability of only a limited number of potential risk factors in the dataset. Further confirmation of the model’s validity and generalizability would require investigation with a broader range of potential risk factors. On the other hand, the current web application is tailored specifically for the analyzed dataset and may have limitations in terms of generalizability to other datasets. To improve usability for medical professionals, it would be valuable to develop a more convenient pipeline that enables users to import and download their own datasets. This enhancement would provide a more flexible and adaptable platform, allowing users to apply the analysis framework to their specific datasets and expand the potential applications of the web application in diverse clinical settings.

## Data Availability

All data produced are available online at

https://easy.dans.knaw.nl/ui/datasets/id/easy-dataset:124398/tab/2

